# Frequency of hospitalization of infants with bronchiolitis during 2017 in Puerto Madryn, Argentina

**DOI:** 10.1101/2021.01.28.21250671

**Authors:** Damián L. Taire, Bruno A. Pazos

**Affiliations:** Departamento de Neumonología Pediátrica, Hospital Zonal “Dr. Andrés R. Isola”, Puerto Madryn U9120, Argentina; Instituto Patagónico de Ciencias Sociales y Humanas, Centro Nacional Patagónico, Consejo Nacional de Investigaciones Científicas y Técnicas, Puerto Madryn U9120, Argentina; Laboratorio de Ciencias de las Imágenes, Departamento de Ingeniería Eléctrica y Computadoras, Universidad Nacional del Sur, Bahía Blanca B8000, Argentina

**Keywords:** respiratory tract infections, socioeconomic factors, epidemiology

## Abstract

**Introduction:** Bronchiolitis is considered the most frequent disease in infants and still represents an important cause of morbidity and mortality worldwide. Despite its viral etiology, socioeconomic variables could influence the disease outcome. We aimed to determine the frequency of hospital discharge for bronchiolitis in a local Hospital in the city of Puerto Madryn, in the province of Chubut, Patagonia Argentina.

**Population and methods:** We performed a cross-sectional study that analyzed all hospitalized patients discharged for bronchiolitis in Hospital “Dr. Andrés R. Isola” during the year 2017 and based on data provided by the hospital administrative staff. The study variables were the length of stay, readmission rate and place of origin of hospitalized patients.

**Results:** A total of 120 patients were included. The median age was 4.45 months (3.9-5). The mean length-of-stay (LOS) was 7.30 days (5.52-9.08). Of the total number of patients, 24 (20%) had a LOS ≤3 days and 96 (80%) a >3 days. One hundred patients (88.33%) had no hospital readmissions and 10 patients (8.33%) had hospital readmissions. The median age of patients with readmissions was 4.2 months (2.69-5.71). The mean LOS during readmission was 17.3 days (5.25-29.35). Of the 120 hospitalized children, 100 infants (83.33%) live in areas identified as having “unsatisfied basic needs” in Puerto Madryn.

**Conclusions:** The overcrowding as a result of the demographic transformation on the frequency of hospitalization of infants with bronchiolitis was homogeneous within the Puerto Madryn population with “unsatisfied basic needs”.

## INTRODUCTION

Acute Lower Respiratory Tract Infections (ALRTI) are one of the main causes of death in the world, with more than 4 million deaths per year (1). To this day, ALRTI still represents an important cause of morbidity and mortality in Argentina (2). In particular, bronchiolitis and pneumonia (with or without complications) are the most relevant because of their impact on morbidity and mortality on child patients (3).

In 2005, ALRTI were responsible for 65,475 hospital discharges on patients under 5 years old in Argentina, representing 21.2% of the total registered for that year (4). The most recent report (29/01/2020) emitted by the National Institute of Respiratory Diseases “Dr. Emilio Coni” concludes that, although the mortality caused by respiratory diseases in children under 5 years old in Argentina continues in a decaying trend, a disparity persists regarding the distribution of mortality due to respiratory diseases in children under 5 years old in Argentina (5). Despite the high number of investigations focused on ALRTI epidemiology around the world, the influence of local factors (geographical, climatic, socioeconomic, cultural) raises the need for every region to have their own registrations and evaluations (6).

The city of Puerto Madryn (Chubut, Northern Patagonia Argentina), where this study was made, has suffered an important demographic transformation between the years 1970 and 2010. Due to an internal and international migration process, the city has multiplied by thirteen its population in the aforementioned period, going from 6.100 to almost 80.000 habitants, being Puerto Madryn the most origin diverse migratory destination in Patagonia (7). The migratory process is a complex phenomenon that involves people (migrants and their families) and societies and has multiple dimensions and consequences such as a rise in overcrowding, that is a known risk factor for respiratory infections.

Puerto Madryn is a coastal city that has transformed in an intermediate size agglomeration due to a high demand for labor from the fishing and construction industries and the expansion of a large aluminium plant located in the outsides of the city (8).

The term bronchiolitis is generally applied to the first episode of infectious sibilances in infants under 12 months of age (9). In addition to the morbidity and mortality caused by it, this pathology demands an important amount of health resources, in particular when hospitalization is required.

At the moment, there is a small amount of information regarding bronchiolitis in Patagonia Argentina. There is an urgent need to establish the relationship between maternal and child morbidity and mortality and poverty in small localities and at an urban level, in the different neighborhoods where the obtained information can be of great use to plan, prioritize, develop, monitor and evaluate the effects of targeted actions to the most vulnerable groups of people living in them. Several authors have mentioned the importance of analyzing neighborhoods in relationship with the health conditions of the people living in them (10) (11) (12) (13).

The objectives of this study are to describe the hospitalization frequency of children younger than 1 year of age with bronchiolitis in Hospital Zonal “Dr. Andrés R. Isola” from Puerto Madryn city during the year 2017 and to explore the use of georeferenced data as a predictor of ALRTI hospitalizations.

## POPULATION AND METHODS

### Design

Cross-sectional study.

### Study inclusion criteria

Children younger than 12 months of age, hospitalized between January 1st, 2017 and December 31st, 2017 with bronchiolitis diagnostic (ICD-10 J21) that have required oxygen therapy

### Study exclusion criteria

Patients that did not require oxygen therapy and those with other concomitant respiratory or cardiological diseases.

### Clinical case definition

Acute Bronchiolitis (ICD-10 J21).

### Indicators

Hospitalization duration or length-of-stay (LOS, expressed in days), readmissions and place of origin of hospitalized patients. Readmissions (total, ≤30 days and >30 days) is the percentage of discharges produced by the same patient in a limited period of time after their index episode. To determine patient places of origin, a Puerto Madryn map was used in conjunction with the Google Maps Geocoding API to translate every patient address in a latitude longitude tuple and be able to locate them accurately on a map. Additionally, this study was made using the geographical codification of homes according to presence or absence of Unsatisfied Basic Needs (UBN) as an overcrowding indicator. The UBN refers to the relationship between the total number of home members and the number of exclusive use rooms in such homes. Operationally, it is considered that critical overcrowding exists in a home when there are more than three people per room (14) (15).

### Data collection instruments

A patient data form with the following fields: birthdate, age in months, admission date, discharge date, address, main discharge diagnostic with its corresponding alphanumeric code according to the International Statistical Classification of Diseases and Related Health Problems, 10th revision (ICD-10: 2016) and oxygen requirement. The data was obtained from the hospitalization archive of the hospital, corresponding to the year 2017. All the gathered information was primarily processed using Microsoft Excel ^®^.

### Ethical considerations

The protocol was approved by a Sub-Committee of Investigation Ethics from the Committee of Bioethics from the Northern Programmatic Area of Chubut province.

### Statistical Analysis

According to a Kolmogorov-Smirnov test, the variables involved could not be adjusted to a normal distribution. As a result, these were summarized as median (interquartile range - IQR). Categorical variables are represented as numbers (percentages) and their 95 % confidence intervals (CI). In this study, we explored the following statistical relationships: the Pearson correlation coefficient between age (months) and length-of-stay (days); the statistical comparison of population subgroups according to the presence or absence of UBN by the asymptotic non parametric Mann-Whitney test, and the statistical relationship between patients living in homes with UBN with readmissions (Chi-square test), age (months) and mean length-of-stay (Mann-Whitney test).

Additionally, subgroups characteristics of patients with and without readmissions were compared using Student’s T-test and Chi-square test accordingly. All the statistical tests used a significance level of *P* < 0.05.

## RESULTS

From the total of infants younger than 12 months of age discharged from the hospital for bronchiolitis (ICD-10 J21) during the study period (n=139), 120 (86.33%) met the inclusion criteria and were selected for the study. From the remaining 19 patients, 18 (94.73%) were excluded due to their discharge diagnostic not being bronchiolitis. The last patient was not included due to an error during the data collection in which its home address was not registered correctly.

The median age (IQR) for the 120 included patients in the study was of 4.45 months of age (3.9-5). The groups distribution by age being: 12 (10%) <1 month old, 76 (63.33%) between 1 and 5 months old, and the remaining 32 (26.66%) between 6 and 11 months old.

The mean (total) length-of-stay was 7.30 days (5.52-9.08). From the total of patients, 24 (20%) had a stay that lasted less or exactly 3 days while 96 (80%) a stay that lasted more than 3 days. The Pearson correlation coefficient is considered positive (*P* = 0.82, > 0.05) (Figure 1). Hospital discharges were granted more frequently on friday and saturday (23 and 24, respectively). The remaining weekdays had the following decreasing trend, tuesday (22), wednesday (20), thursday (11), monday and sunday (10).

**Figure 1.**
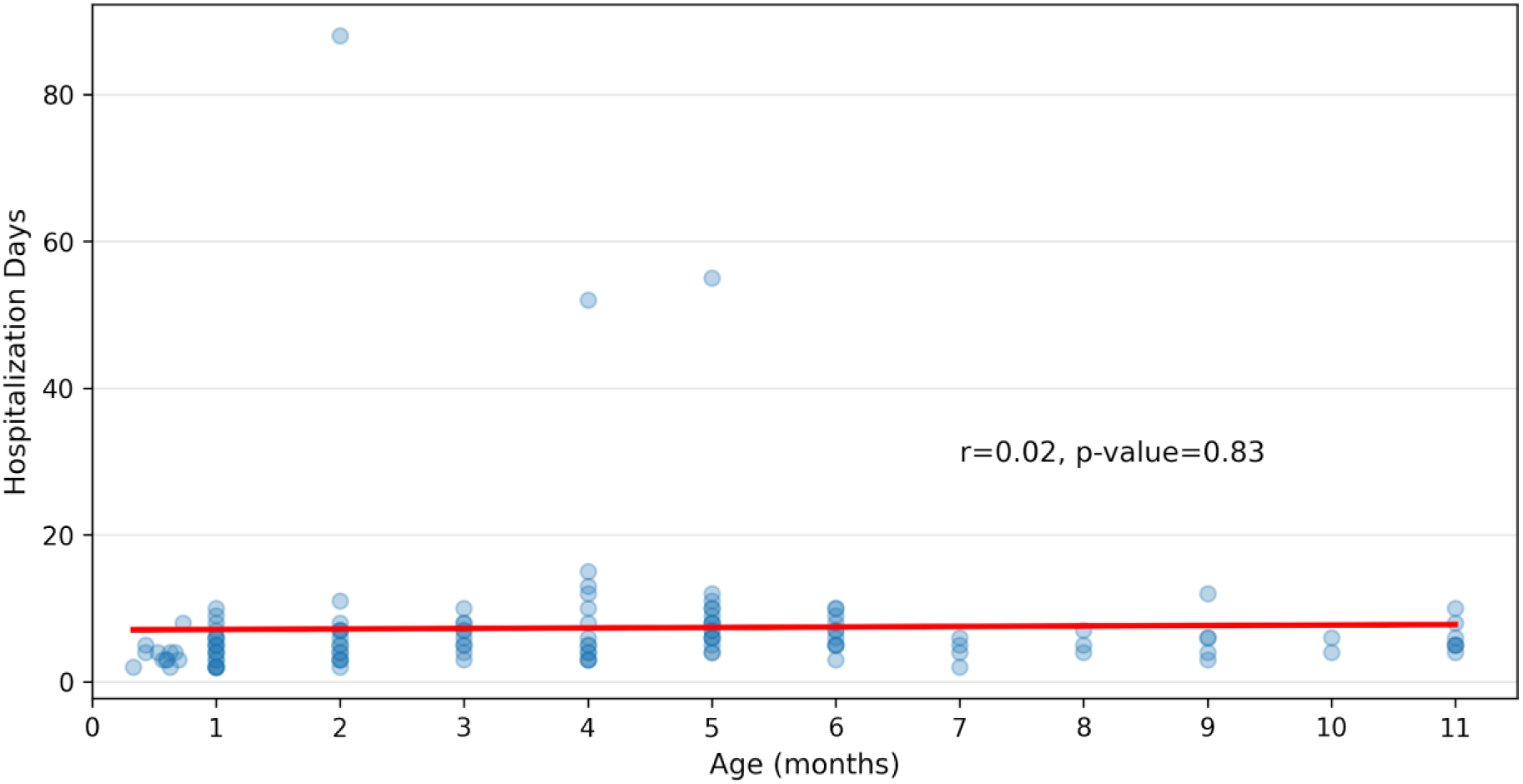
Age (months) vs. Hospitalization days.

Between april and october of 2017, 102 patients (85%) were hospitalized with a seasonal peak of epidemiological weeks (EW) in the range of 28 and 31, corresponding to the month of july (Figure 2) from the same year. One of the surveyed indicators was the place of origin of the hospitalized patients (districts, sector, neighborhoods and number of blocks), illustrated on the city map (Figures 3 and 4). Of the 120 hospitalized patients, 100 (83.33%) lived in sectors corresponding (mainly) to homes registered as with the presence of UBN during both Argentina’s 2001 and 2010 national population and housing surveys. The mean age of the 100 infants was 4.31 months of age (1.51-7.11). The groups distribution by age being: 76 (76%) younger than 6 months of age and 24 (24%) older than 6 months of age. The length of stay had a mean of 7.57 days in total (5.46-9.68).

**Figure 2.**
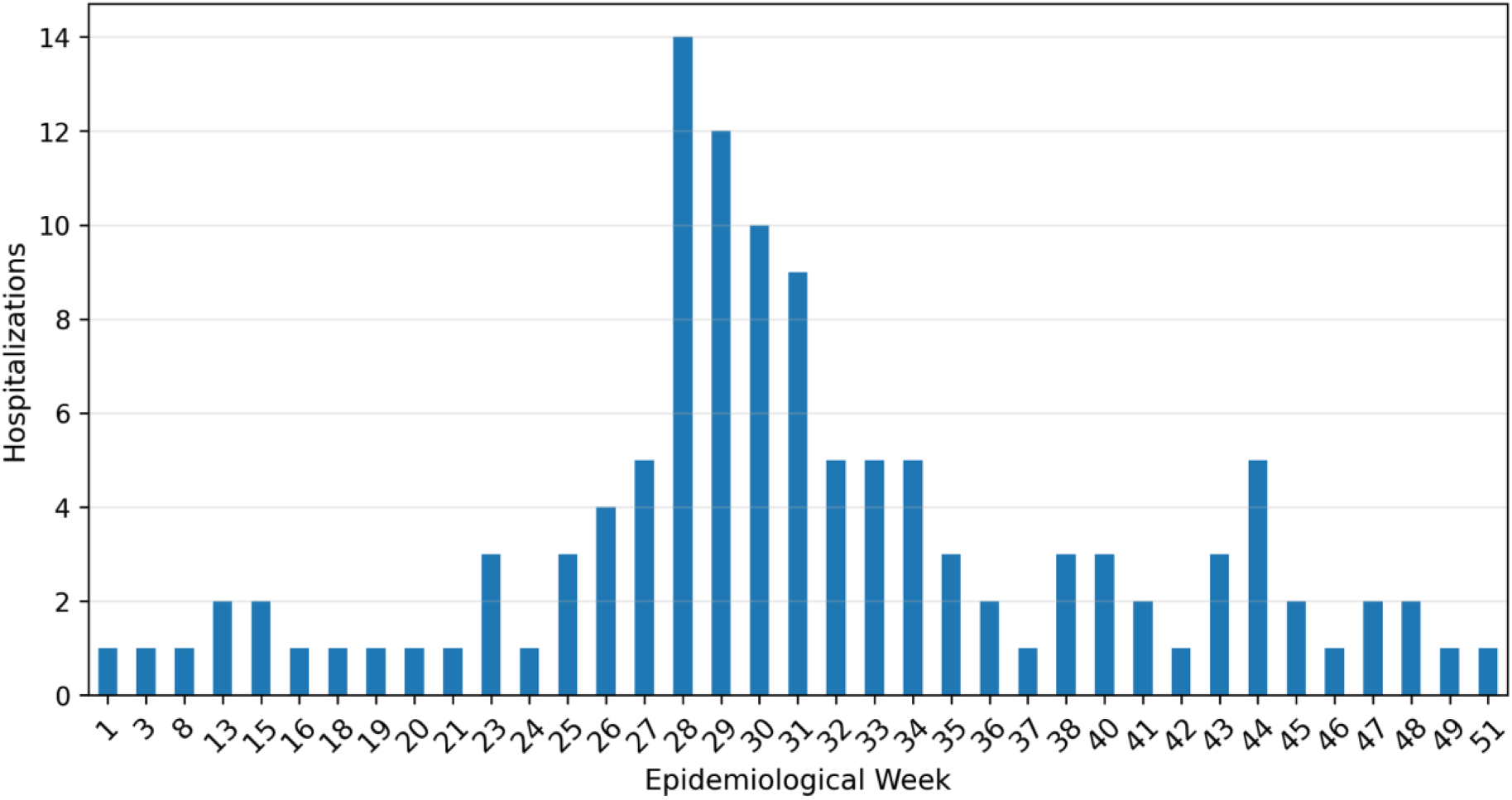
Pediatric hospitalizations for Bronchiolitis, according to Epidemiological Week.

**Figure 3.**
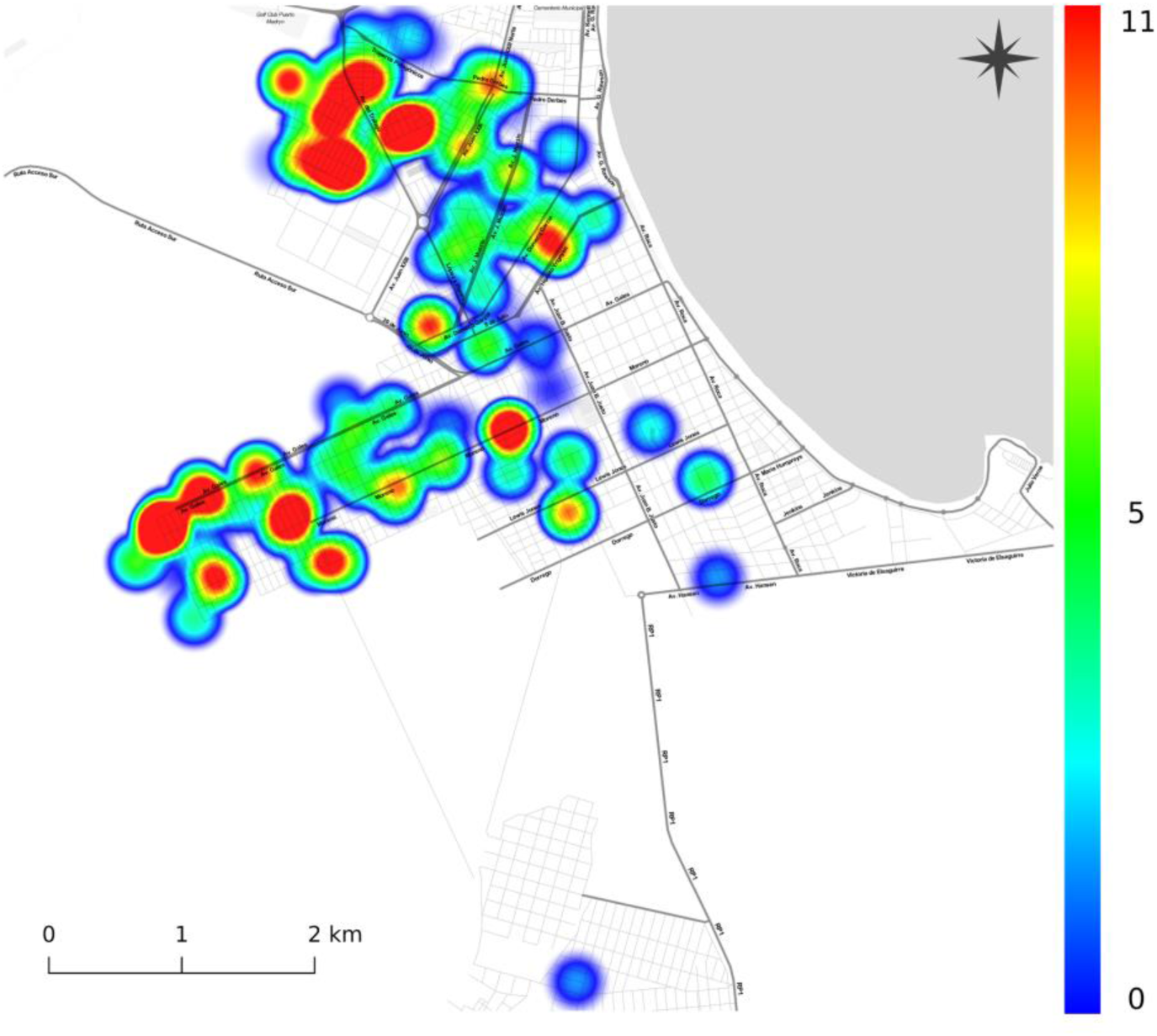
Heatmap, Age (months) and patient home address.

**Figure 4.**
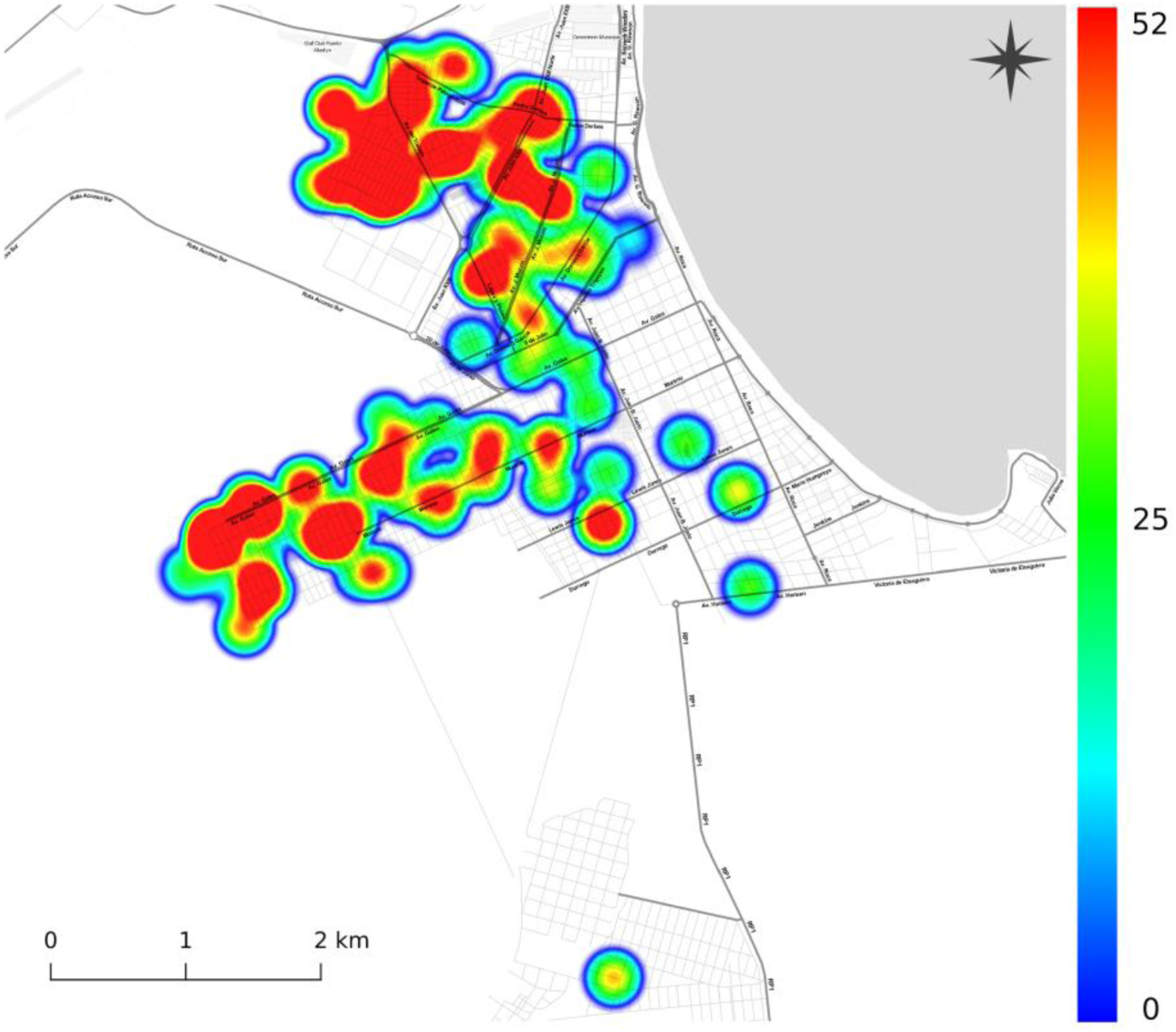
Heatmap, Epidemiological Weeks and patient home address.

Regarding the number of readmissions by bronchiolitis during the period of study, 110 patients (91,66%) presented no need for readmission while 10 of them (8,33%) needed to be readmitted.

From the latter group, 7 were readmitted in a period ≤ 30 days after their index episode and the remaining 3 in a period > 30 days after their index episode. The median age (IQR) of the readmitted infants was 4.2 months of age (2.69-5.71), 3.71 months of age (1.62-5.79) for those readmitted ≤ 30 days. There was no significant difference observed for the ages of the subgroups with and without readmissions (*P* = 0.50). The mean length-of-stay of the readmissions was 17.3 days in total (5.25-29.35). There also was no significant difference found between hospitalization days for the subgroups with and without readmissions (*P* = 0.99). 9 out of 10 readmissions were from patients living in sectors corresponding to those with homes in the presence of UBN according to Argentina’s 2001 and 2010 national population and housing surveys. No statistical relationship was found between subgroups with and without readmissions regarding the presence or absence of UBN (Chi-squared = 0.18). The detailed frequencies of patients with admissions and readmissions related to bronchiolitis can be seen in Table 1.

**Table 1.**
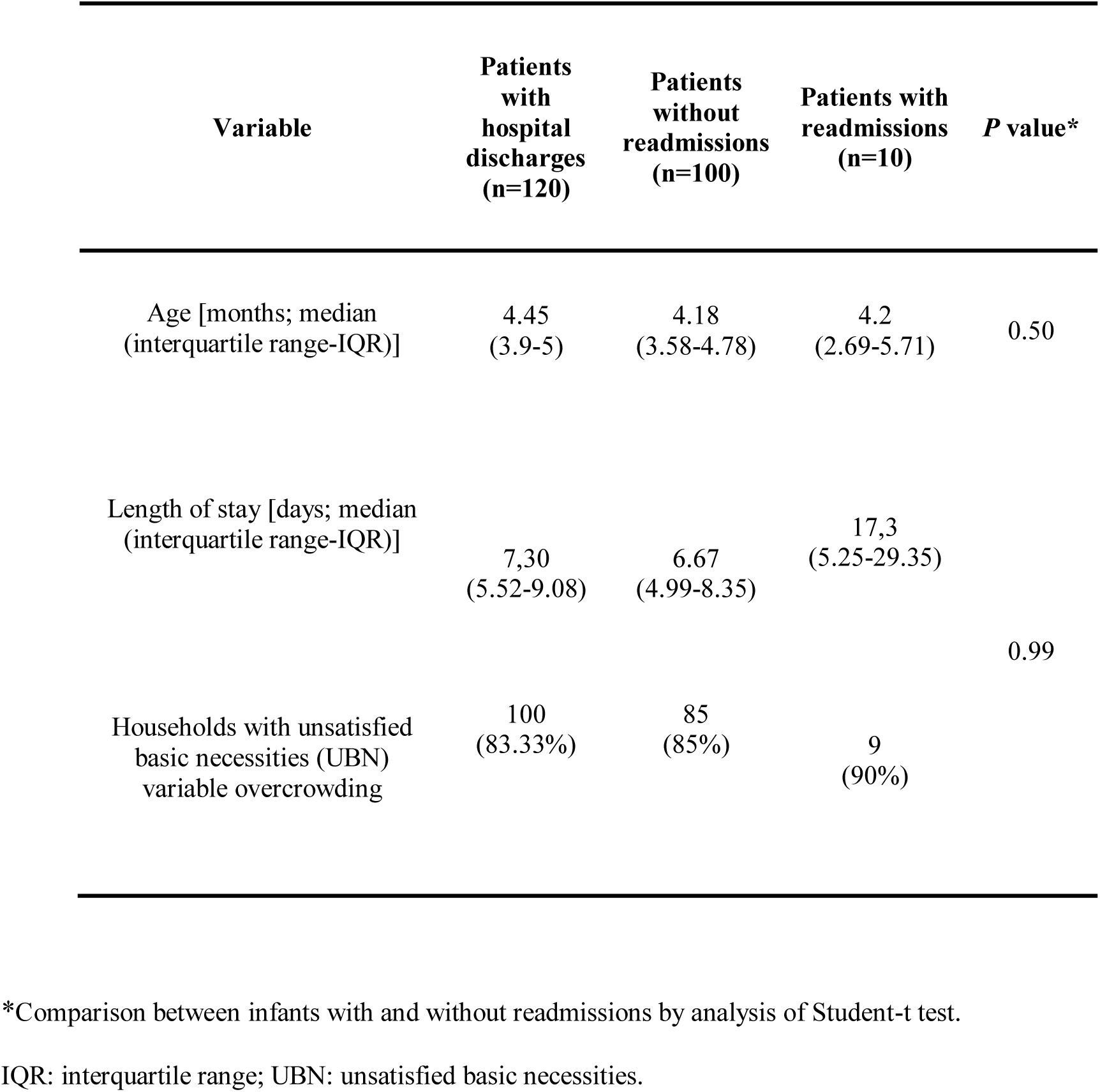
Hospital discharges and readmissions for bronchiolitis during the study period.

| Variable | Patients with hospital discharges (n=120) | Patients without readmissions (n=100) | Patients with readmissions (n=10) | P value* |
| --- | --- | --- | --- | --- |
| Age [months; median (interquartile range-IQR)] | 4.45<br>(3.9-5) | 4.18<br>(3.58-4.78) | 4.2<br>(2.69-5.71) | 0.50 |
| Length of stay [days; median (interquartile range-IQR)] | 7,30<br>(5.52-9.08) | 6.67<br>(4.99-8.35) | 17,3<br>(5.25-29.35) | 0.99 |
| Households with unsatisfied basic necessities (UBN) variable overcrowding | 100<br>(83.33%) | 85<br>(85%) | 9<br>(90%) |  |

\*Comparison between infants with and without readmissions by analysis of Student-t test.
IQR: interquartile range; UBN: unsatisfied basic necessities.

## DISCUSSION

This study found that a high proportion (83.33%) of infants hospitalized for bronchiolitis during the year 2017 in Puerto Madryn, lived in sectors corresponding to homes in the presence of UBN. This method has the advantage that allows the elaboration of poverty maps at a district level when national population and housing surveys are used as a source of information (16). As an example, based on the 2010 survey, the National Institute of Statistics and Surveys elaborated cartography based on the information provided by provincial statistical institutes. At the time, 9.42% of the total population from Puerto Madryn lived in homes in the presence of UBN (17).

Of all the factors that make up a deficit situation, overcrowding, in multiple authors’ opinion, is the most serious indicator of the deficit due to the wide range of negative consequences it causes. Infants are particularly vulnerable to this situation (18) (19). Overcrowding rises the number of family members cohabiting that are able to spread respiratory infections to the babies.

Precarious housing conditions include sanitation problems (not having access to mains water inside the home, not having a flush toilet and/or not having access to the sewer network) (19), access to gas and electricity supply networks, minimum thermal insulation and minimum ventilation/person.

Current findings may have important consequences to predict future hospitalizations by bronchiolitis in infants given the fact that, for what it’s known, this is the first study made in Patagonia Argentina according to the place of origin of the infants, that looks for bronchiolitis hospitalization predictors without any intervention.

In the present patagonian study, a seasonal peak between the EW 28 and 31 was evidenced, corresponding to the month of july of 2017. The bronchiolitis notification curve in children under 2 years old in Argentina during 2017 stayed within the safety and success zone, reaching a seasonal peak during EW 24 (20).

One of the strengths of this study with a hospitalary base, is that it allows to infer results for the general population of Puerto Madryn, being this population representative of homes in the presence of UBN. The database was designed and developed by administrative personnel specialized in hospital auditing and supervised by one of the authors (DLT), which generates acceptable confidence in the collection and recording of the data.

Related factors (access to health services, network infrastructure and equipment, health specialist human resources, etc) and probability or readmission were not evaluated due to the lack of information of outpatients after their first documented discharge (21) (22). In this study there was no information on the main diagnostic of patients readmission, so it is unknown whether they were readmitted for reasons other than bronchiolitis (23).

All the generated information was georeferenced allowing the creation of maps that facilitate the task of visualizing problematic areas for the public health authorities and local governments to line up resources to optimize the maternal and child health system complementing each other in the process.

## CONCLUSION

The process of overcrowding as a result of a demographic transformation on the frequency of infant hospitalization due to bronchiolitis was homogeneous within the Puerto Madryn population given the fact that the majority of it lives in homes in the presence of UBN. The challenge continues to be transforming the knowledge of the populations we study towards the development and execution of more effective health interventions.

## Data Availability

All the aggregate data generated and analysed during the current study are included in this published article. The data sets are available from the corresponding author upon request.

## Author Contributions

D.L.T. conceived the original idea. D.L.T. made the clinical data collection.

D.L.T. and B.A.P. analyzed the results. B.A.P. developed the distribution plots and heatmaps. D.L.T. wrote the manuscript. All authors revised and agreed to submit the current version of the manuscript.

## Acknowledgments

Authors would like to thank Dr. Fernando C. Ferrero, who kindly review the earlier version of this manuscript and provided valuable suggestions and comments; and to the administrative staff of the Puerto Madryn Hospital for the collaboration in the registration of the data necessary for this research during 2017.

## Abbreviations

The following abbreviations are used in this manuscript:

LOS: length-of-stay
ALRTI: acute lower respiratory tract infections
ICD-10: International Classification of Diseases and Related Health Problems, tenth revision
IQR: interquartile range
CI: confidence interval
UBN: unsatisfied basic necessities
EW: epidemiological week

